# Characterizing Treatment Non-responders vs. Responders in Completed Alzheimer’s Disease Clinical Trials

**DOI:** 10.1101/2023.10.27.23297685

**Authors:** Dulin Wang, Yaobin Ling, Kristofer Harris, Paul E. Schulz, Xiaoqian Jiang, Yejin Kim

## Abstract

Alzheimer’s disease (AD) patients have varying responses to AD drugs and there may be no single treatment for all AD patients. Trial after trial shows that identifying non-responsive and responsive subgroups and their corresponding moderators will provide better insights into subject selection and interpretation in future clinical trials. We aim to extensively investigate pre-treatment features that moderate treatment effect of Galantamine, Bapineuzumab, and Semagacestat from completed trial data. We obtained individual-level patient data from ten randomized clinical trials. Six Galantamine trials and two Bapineuzumab trials were from Yale University Open Data Access Project and two Semagacestat trials were from the Center for Global Clinical Research Data. We included a total of 10,948 subjects. The trials were conducted worldwide from 2001 to 2012. We estimated treatment effect using causal forest modeling on each trial. Finally, we identified important pre-treatment features that determine treatment efficacy and identified responsive or nonresponsive subgroups. As a result, patient’s pre-treatment conditions that determined the treatment efficacy of Galantamine differed by dementia stages, but we consistently observed that non-responders in Galantamine trials had lower BMI (25 vs 28, *P* < .001) and increased ages (74 vs 68, *P* < .001). Responders in Bapineuzumab and Semagacestat trials had lower Aβ_42_levels (6.41 vs 6.53 pg/ml, *P* < .001) and smaller whole brain volumes (983.13 vs 1052.78 ml, *P* < .001). 6 ‘positive’ treatment trials had subsets of patients who had, in fact, not responded. 4 “negative” treatment trials had subsets of patients who had, in fact, responded. This study suggests that analyzing heterogeneity in treatment effects in “positive” or “negative” trials may be a very powerful tool for identifying distinct subgroups that are responsive to treatments, which may significantly benefit future clinical trial design and interpretation.

## Introduction

The current understanding and treatment approach towards Alzheimer’s disease (AD) presents substantial challenges, especially in the context of differential responses to medication between patients. Responsiveness (high efficacy) or non-responsiveness (low efficacy) to AD treatment might be related to certain moderators. A previous study indicated that one-third of AD patients do not respond to pharmacological treatment.^1^ Different moderators, such as sex or gender,^2^ the presence of comorbid illness,^3^ and long-term administration of other medications (e.g., antihypertensives),^3^ might contribute to different responses of AD patients to treatments. Because a one-size-fits-all treatment is not likely to be available for AD in the near future, identifying non-responsive or responsive subgroups and their corresponding moderators will provide better insights into subject selection in future clinical trials.

Identifying patient subgroups with differential responses to treatment requires counterfactual analysis, which compares the outcomes of patients who actually received the treatment (factual outcome) with the outcomes when the same patients would not have received the treatment (counterfactual outcome), and vice versa.^4,5^ Counterfactual outcomes are never observed. To estimate the average treatment effect (ATE), the traditional approach takes the difference of potential outcomes between the randomized (or computationally debiased) treatment and placebo arms.^6^ To estimate heterogeneous treatment effect (HTE), earlier studies stratify the samples by prespecified variables and test the statistical significance of differences in arms.^7^

It is also common practice to try to identify subsets of responsive patients in Phase II clinical trials to determine enrollment criteria for the associated Phase III trial. Many Phase III trials following positive Phase II subgroups, however, have shown negative outcomes, including tarenflurbil, bapineuzumab, solanezumab, and ELND005.^8–11^ Therefore, identifying subgroups with differential responses may be beyond the scope of the ad-hoc analyses of a few manually selected variables in individual trials, considering the complexity of AD.^12–16^ Several recent HTE estimation models based on machine learning have flexibly incorporated high-dimensional features space in randomized data. Among them, tree-based HTE estimation models recursively partition subjects in randomized trials to maximize the separation in the subject’s HTE.^17,18^ The model’s ability to account for nonlinearity in pre-treatment features and interpretability motivated us to leverage this approach to estimate HTE and obtain important features that moderate the treatment effect size.

The goal of this study was to identify important pre-treatment features that define subgroups within randomized clinical trials with positive or negative treatment efficacy. For trials with positive efficacy (e.g., Galantamine), we focused on identifying nonresponsive subgroups who are less likely to benefit from the drug in order to not expose those patients to drugs to which they are unlikely to respond. For trials with neutral efficacy (e.g., Bapineuzumab, Semagacestat), we focused on identifying responsive subgroups that might be amenable to targeted trials.

## Methods

### Study population

We obtained individual-level patient data from ten randomized clinical trials (RCTs) on AD drugs from the Yale University Open Data Access Project (YODA, http://yoda.yale.edu/) and Center for Global Clinical Research Data (Vivli, https://vivli.org/). We summarized the ten RCTs in Table 1. The datasets consist of six Galantamine trials for patients with mild to severe AD, two Bapineuzumab trials for mild to moderate AD, with or without ApoE4, and two Semagacestat trials for mild to moderate AD without depression. The Galantamine and Bapineuzumab trials were from YODA and the Semagacestat trials were from Vivli. The primary results of these studies have been previously published.^9,19–22^ Galantamine improved cognitive function but did not significantly enhance overall daily activities. In contrast, both Bapineuzumab and Semagacestat showed neutral efficacy in improving cognitive function, and patients receiving high doses of Semagacestat showed worse outcomes.

**Table 1.**
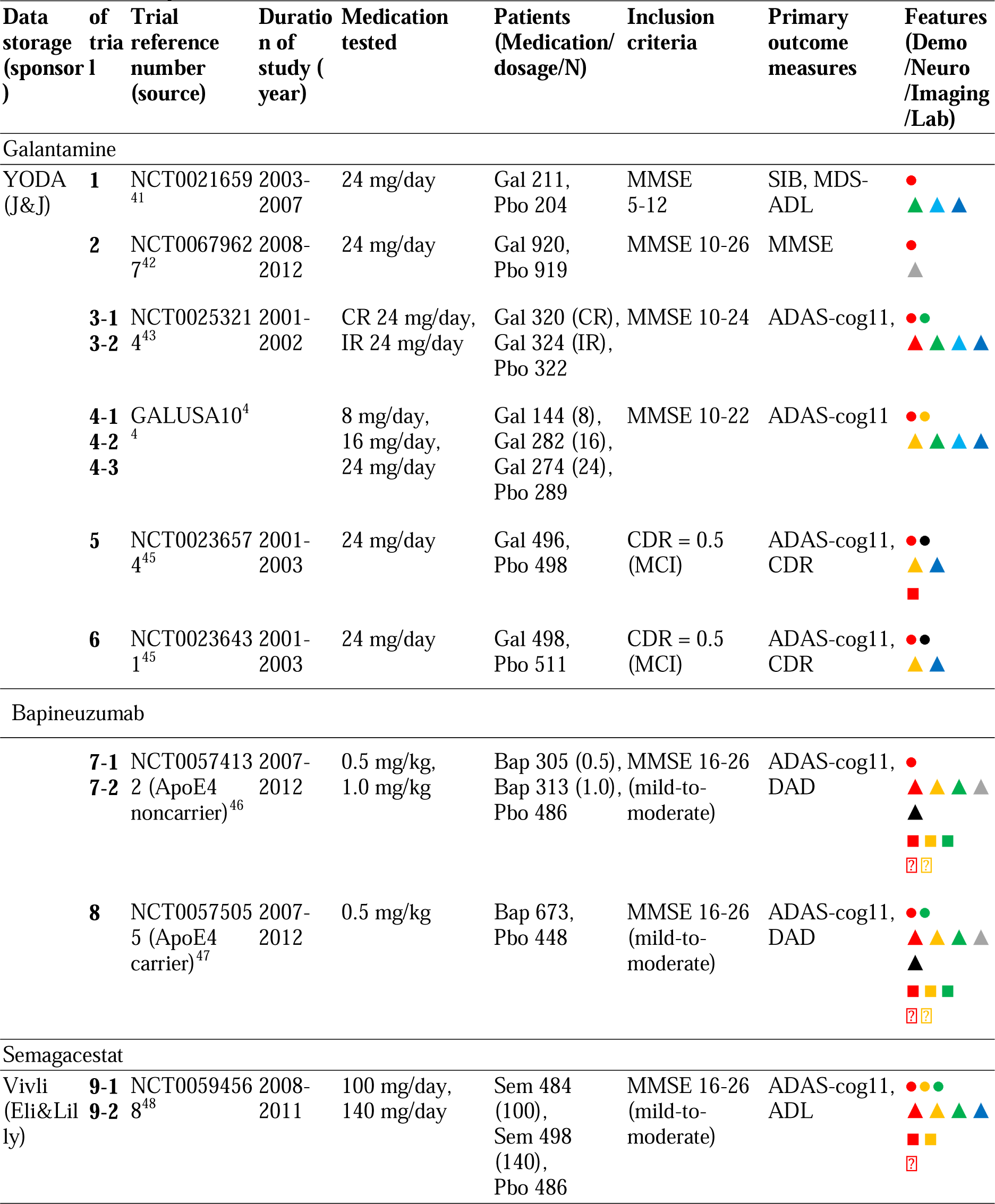
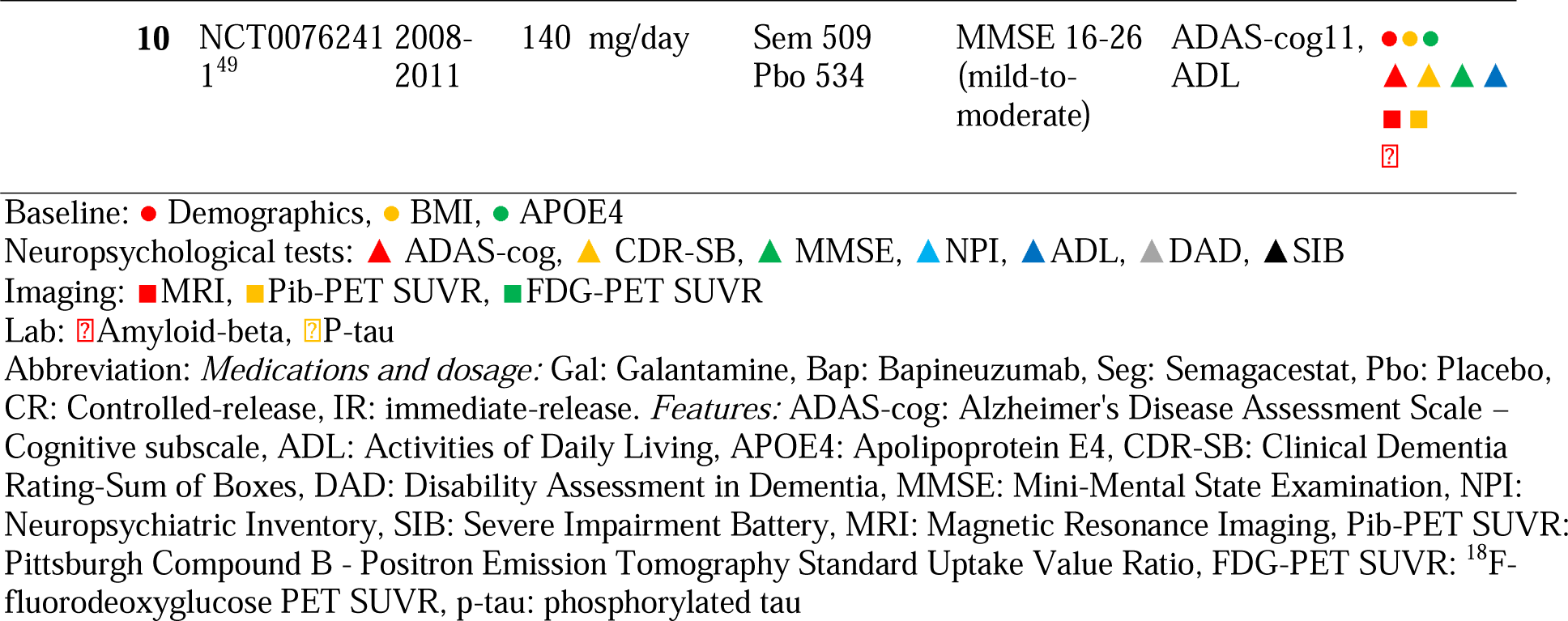
Summary of ten randomized clinical trials.

### Outcome measurement

The ten RCTs use different sets of outcome measurements to evaluate cognitive function or activities of daily living. The outcomes were assessed before treatment (baseline) and after treatment (endpoint). The ten RCTs also have different study durations. The Alzheimer’s Disease Assessment Scale –Cognitive subscale (ADAS-cog) was the primary outcome measure in most RCTs (eight out of ten studies) (Table 1). Our study focused on the most common outcome measurements, which included ADAS-cog, Severe Impairment Battery (SIB), and Mini-Mental State Examination (MMSE), to ensure comparability between trials. Although they measure cognitive function with different tasks, they are developed to evaluate the cognitive function of AD (or prodromal AD) patients, making it possible for us to compare the efficacy across different trials.

### Features

We included a comprehensive set of pre-treatment features to identify features that moderate the treatment effect (moderator). The features include demographic variables, dependencies, neuropsychological tests, Cerebrospinal Fluid Phospho (CSF) biomarkers, and brain imaging. The availability of such features varied by trial (Table 1). These features may moderate the treatment effects. Previous studies have shown that patient demographics, such as age and ethnicity, can impact cognitive function in AD patients.^23,24^ Dependency tests included Activities of Daily Living (ADL) and Disability Assessment For Dementia (DAD). Cognitive tests such as the Clinical Dementia Rating-Sum of Boxes (CDR) and MMSE have commonly been used to evaluate AD patients’ cognitive function.^25^ CSF Biomarkers such as amyloid-beta (Aβ) level and phosphorylated tau (p-tau) have been shown to be useful in detecting AD pathology and predicting cognitive decline.^26^ Imaging studies have shown that regional brain volume changes are associated with cognitive decline and amyloid-beta burden.^27,28^ We scaled the regional brain volume (i.e., hippocampal volume and ventricular volume) by whole brain volume as a representation of the proportion of the brain. We also included Pittsburgh Compound B (PiB) - Positron Emission Tomography Standard Uptake Value Ratio (PET SUVR) and ^18^F-fluorodeoxyglucose (FDG) - PET SUVR as imaging features.

### Missing value imputation

In our study, missing values exist in the imaging features (e.g., whole brain volume, hippocampus volume, ventricular volume, PiB-PET SUVR, and FDG-PET SUVR) and lab test features (e.g., Aβ_42_, Aβ_40,_ and p-tau), with missing rates ranging from 48% to 93%. We dropped missing variables with a missing rate larger than 90%. Other features had missing rates under 10%. To overcome such severe missing rates, we imputed missing values with multiple imputations by chained equations (MICE)^29^. MICE is a valid method to iteratively impute missing values in each variable using regression models that take into account the relationships between those variables and other observed variables. This process is repeated multiple times, with each iteration improving the imputations by incorporating information from the imputed values of other variables. The final imputed dataset is created by combining the results from multiple imputations, which produce a set of plausible values that account for the uncertainty in the imputed values.

### Causal forest

We leveraged causal forest^30^ to estimate conditional average treatment effect (CATE), identified potential moderators that affect the efficacy, and used the moderators to define responders (for neutral trial) and non-responders (for positive trial). Compared to other causal inference methods^31^, causal forests recursively partition the subjects, thus accounting for the non-linear relationships between features and treatment effect size. The causal forest model is an ensemble of causal trees, where causal trees recursively partition subjects into small subgroups, like classical decision trees, but to maximize the distribution divergence of treatment effects across leaf nodes. The technical details on potential outcome framework, causal forest modeling, and training process were depicted in the Supplement Text.

### Important features

We assessed feature importance across all trees in the forest based on their relative position in a tree and influence on CATE. We calculated the feature importance by jointly considering the location of the feature used in a tree and the times that the features selected as the splitting criteria when growing a tree, which is

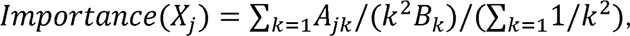

here *k* is the depth of the tree, *A_jk_* is the total number of splits of the variable *X_j_* at depth *k, j* is an index of features included in the algorithm, and *B_k_* is the total number of splits at depth k over every tree in the forest. In addition, to visually investigate the impact and direction of important features as potential moderators on the CATE, we plot the relationships between the top important features and the estimated CATE. We estimated the CATE here from the trained causal forest for each feature value while keeping all the other covariates at the median value.

### Responder and non-responder identification

Traditionally, the responders or non-responders refer to the individuals who have estimated individual treatment effect below zero or above zero. In this study, we presented subgroups of responders or non-responders by selecting leaf nodes in causal trees in the causal forest, where the patients in the subgroups share similar estimated treatment effects and similar pre-treatment features. We focused on leaf nodes that involved important features that we identified. The leaf node is defined as a series of splitting rules and its CATE. In the galantamine trials, which had positive efficacy, we identified nonresponder subgroups. For bapineuzumab and Semagacestat trials, which had negative outcomes, we identified responder subgroups who may have benefitted from the drugs. The higher value of outcome measurement (e.g., ADAS-cog11) means severe cognitive dysfunction, and SIB and MMSE were negated to make the favorable outcome in the same direction as ADAS-cog11. Thus, negative treatment effects indicate beneficial treatment effects. In addition, we tested the important features’ mean difference between subgroups with ITE above or below zero to validate the observed patterns using a two-sample t-test.

### Evaluation

To evaluate whether the causal forest accurately estimated the CATE, we applied the treatment heterogeneity test by analyzing the “best linear predictor” ^32,33^. In the test, we fitted the estimated CATE as a linear function of out-of-bag causal forest estimates. The function has two synthetic regressors: mean forest prediction accounts for the average of out-of-bag CATE estimates with a coefficient a, and differential forest prediction measures the quality of the estimates of treatment heterogeneity on out-of-bag data, with a coefficient β^33^. The coefficients α and β allow us to evaluate the performance of our estimates. If α = 1, then the average prediction produced by the forest is correct. Meanwhile, if β = 1, then the forest predictions adequately capture the underlying heterogeneity. For tests with β close to 0 or negative, the test between above and below the median of the overall range of the CATE provides us with qualitative insights about the strength of heterogeneity.

## RESULTS

Our study included six Galantamine trials, two Bapineuzumab trials, and two Semagacestat trials. Trial details can be found in Table 1.

### Non-responders vs. Responders

We built causal forests on ten trials and characterized responders and non-responders. The estimated CATE for patients (Figure 1) in Trial 1 to 4 were all below 0, indicating that patients were all responders when looking into individualized CATE. It is worth mentioning that we can still find non-responsive subgroups with positive estimated CATE by extracting splitting rules from causal trees. To simplify, the estimated CATE refers to the subgroup CATE. Table 2 summarizes the important features from causal forests and characteristics of non-responders (in Galantamine trials) or responders (in Bapineuzumab and Semagacestat trials) obtained from causal trees, to identify common patterns in non-responders or responders across trials. In the following, we will discuss the findings by drug.

**Figure 1.**
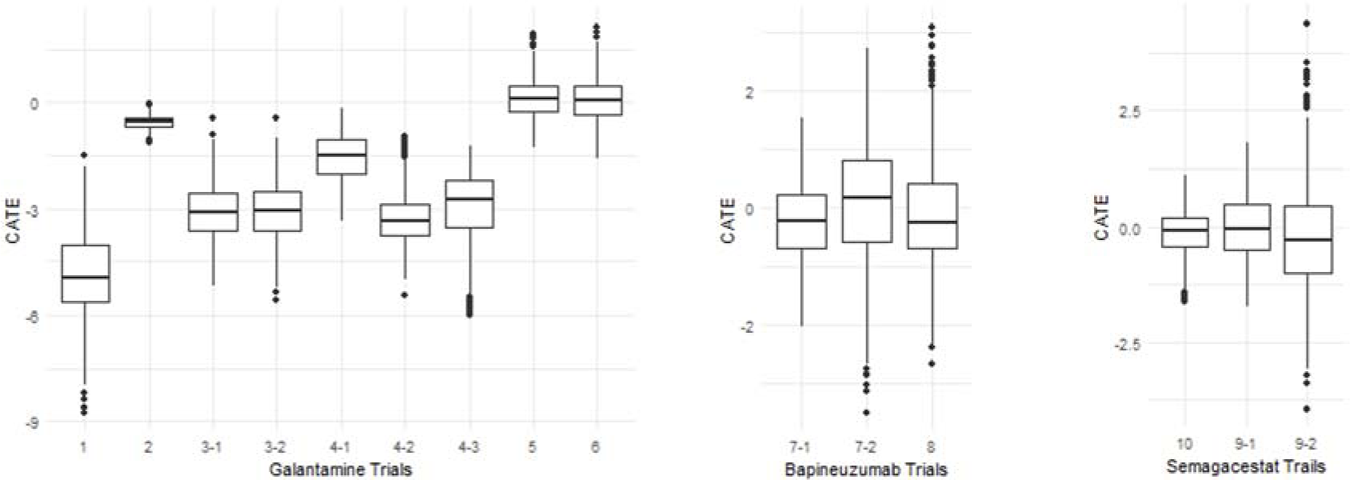
Estimated CATE across ten trials.

**Table 2.**
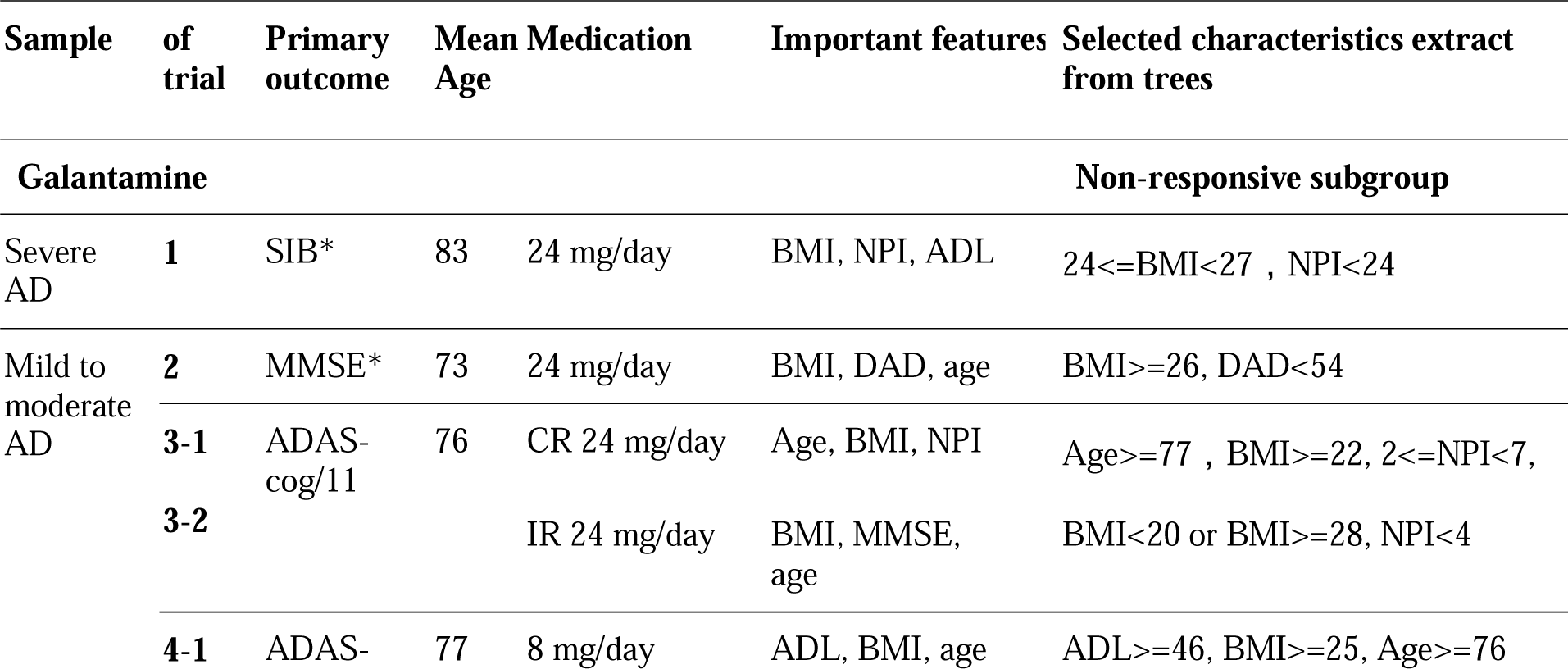
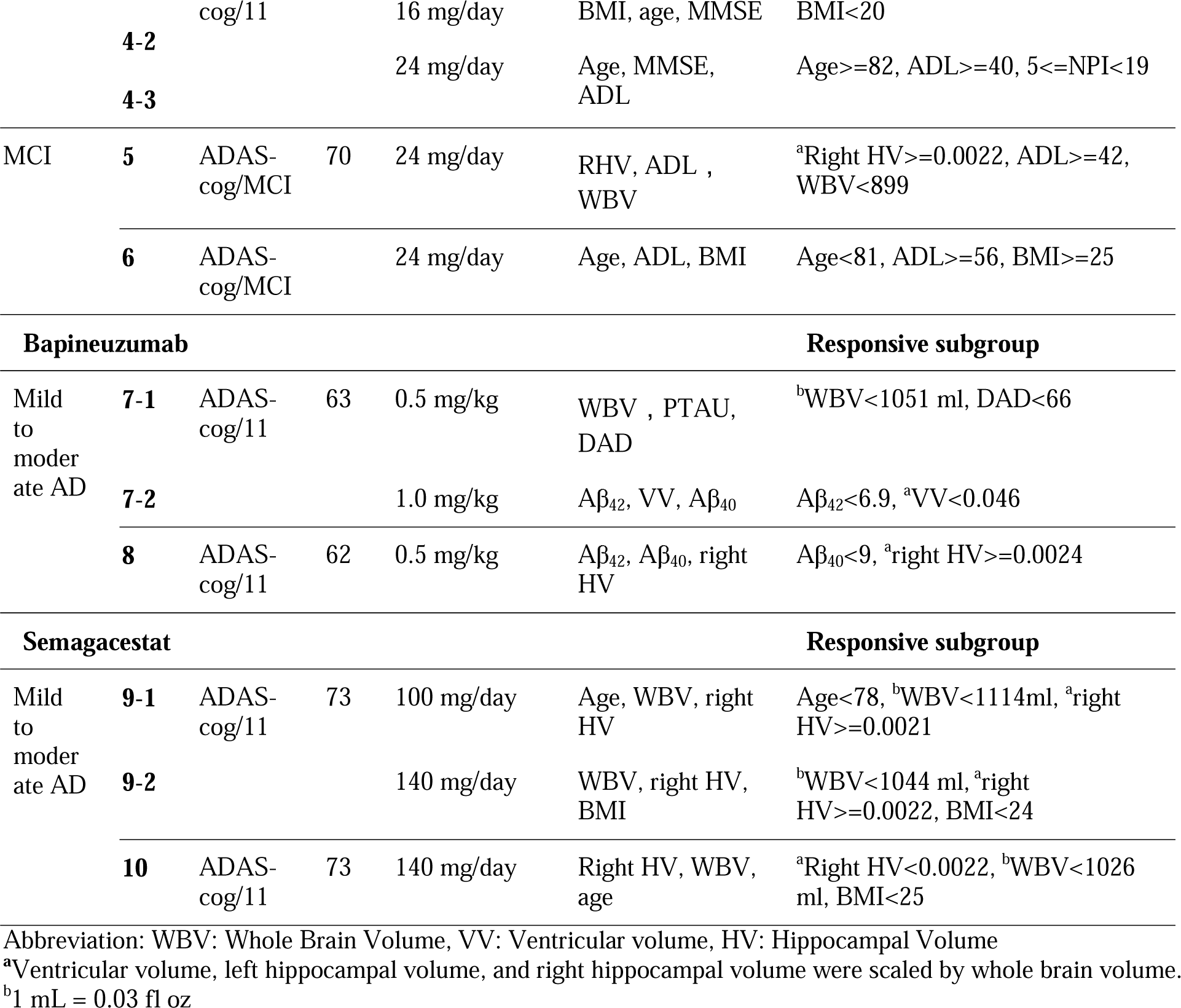
Characteristics of non-responsive and responsive subgroups using the causal forest.

BMI was a recurring important feature across the different stages of dementia and different trials. Among severe AD patients, non-responders typically had a BMI between 24 and 27 and NPI total scores below 24. In contrast, non-responders in the mild to moderate AD patients in trials 2-4 presented with various characteristics. Commonly observed were individuals who were overweight (BMI>=25), had higher ADL total scores (>=46; range from 0 to 78, with a lower score indicating greater severity), and relatively lower NPI total scores range (e.g., [2,7), (0,4), and [5,19); range from 0 to 144, with a higher score indicating greater severity). We also observed that the underweight (BMI<20) group did not respond to the Galantamine in trials 3 and 4.

Brain volume metrics were also pivotal factors influencing treatment response. In trial 5, MCI non-responders tended to have increased right hippocampal volume (>=0.0022) and reduced whole-brain volume (<900 ml). For MCI patients in trial 6, non-responders were overweight (BMI>=25) and younger (age<81). Both non-responder subgroups in trials 5 and 6 had higher ADL total scores (<=42). We further investigated the distinction in selected features between patients with responsive and non-responsive subgroups (eTable 1 in the Supplement). Since only trials 5 and 6 show heterogenous ITE, we only focused on the feature distinction in these trials. In trial 5, we can observe the significant feature distinction in whole brain volume (1092.54 vs. 1060.67 ml, *P* < .001) and right hippocampal volume (2.43e-3 vs. 2.79e-3, *P* < .001). In trial 6, we can observe the significant feature distinction in age (74 vs. 68, *P* < .001), ADL total scores (48 vs. 53, *P* < .001), and BMI (25 vs. 28, *P* < .001).

Across Bapineuzumab trials, mild to moderate AD patients who responded favorably to the treatment exhibited lower Aβ_42_ (<6.9) levels in trial 7 with high-dose treatment and lower Aβ_40_ (<9) levels in trial 8. Both Aβ_42_ (6.41 vs. 6.53 pg/ml, *P* < .001) and Aβ_40_ (8.64 vs. 8.95 pg/ml, *P* < .001) are significantly different between subgroups with negative ITE and positive ITE. In addition, the responsiveness was influenced by brain volume metrics in each trial. These features were also significantly different in negative and positive ITE subgroups. In trial 7, responders had smaller whole brain volume (<1051 ml; 983.13 vs. 1052.78 ml, *P* < .001) with low-dose treatment and smaller ventricular volume (<0.046; 0.045 vs. 0.056, *P* < .001) with high-dose treatment. In trial 8, responders had larger right hippocampus volume (>=0.0024, 2.28e-3 vs. 2.13e-3, *P* < .001).

In the Semagacestat trials, the whole brain volume and right hippocampal volume were consistently observed in the characteristics of responders. In trial 9, responders in two treatment groups typically exhibited smaller whole brain volume (<1114ml or <1044 ml) and larger right hippocampal volume (>=0.0022). There was a significant difference in whole brain volume between negative and positive responsive subgroups (low-dose group: 1062.25 vs. 992.07 ml, *P* < .001; high-dose group: 1014.41 vs. 1037.06, *P* = .003). The right hippocampal volume only showed differences in the low-dose group (1.87e-3 vs. 1.64e-3, *P* < .001). In contrast, in trial 10, responders had smaller whole brain volume (<1044 ml, 967.53 vs 1057.65, *P* < .001) and smaller right hippocampal volume (<0.0022, 1.73e-3 vs. 1.70e-3, *P* = 0.02). Demographic characteristics also played a significant role. Responders in trial 9 treated with low-dose tended to be younger (age<78; 69 vs. 78, *P* < .001), and responders in trial 10 maintained a BMI within the normal range (BMI<25; 24 vs. 25, *P* < .001).

### Relationship between CATE and moderators

We further investigated the potential moderators affecting the CATE estimation by analyzing the impact of single feature alterations on the CATE. Graphical depictions of the relationships between prominent features and the estimated CATE were provided in Figure 2.

**Figure 2.**
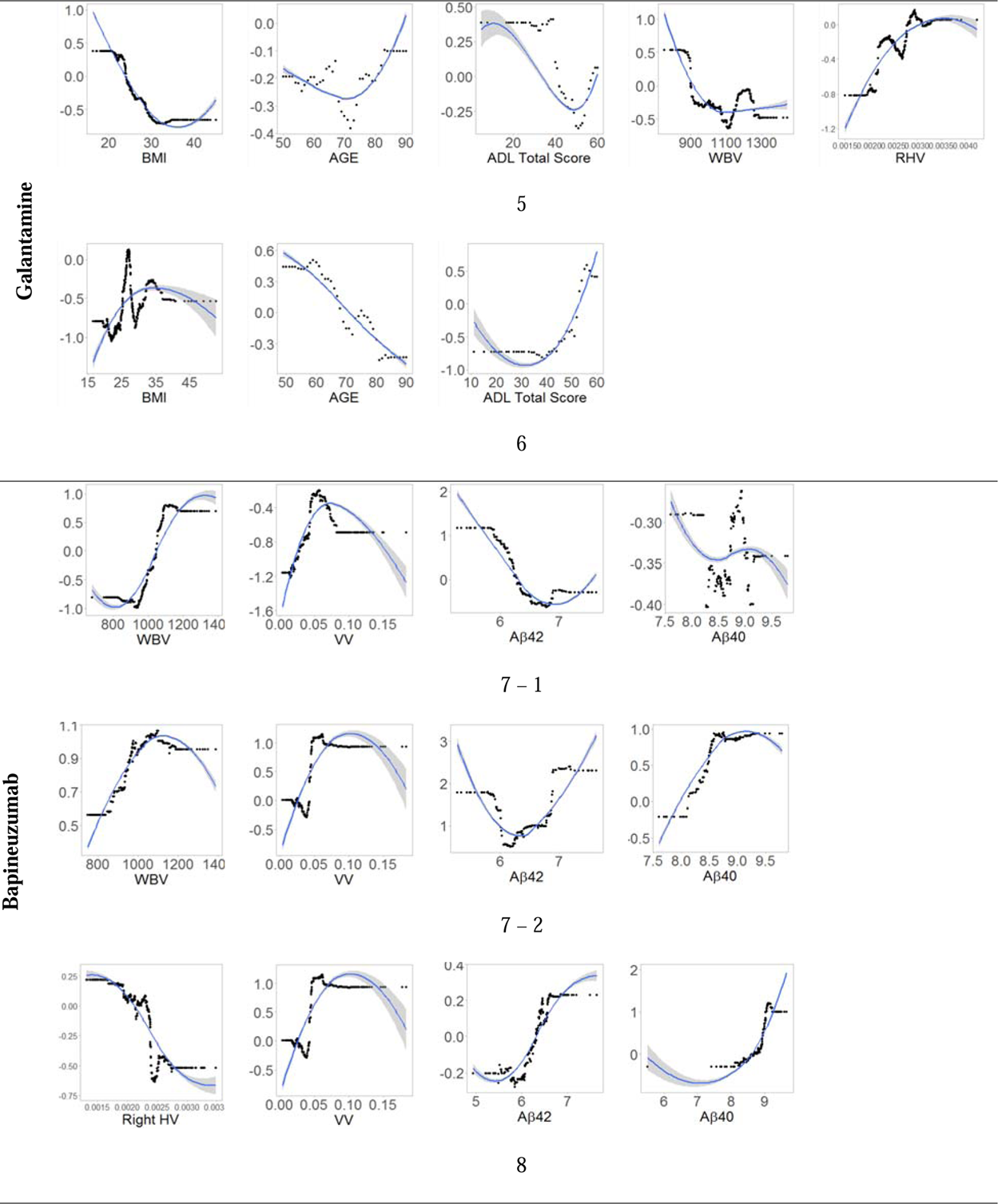
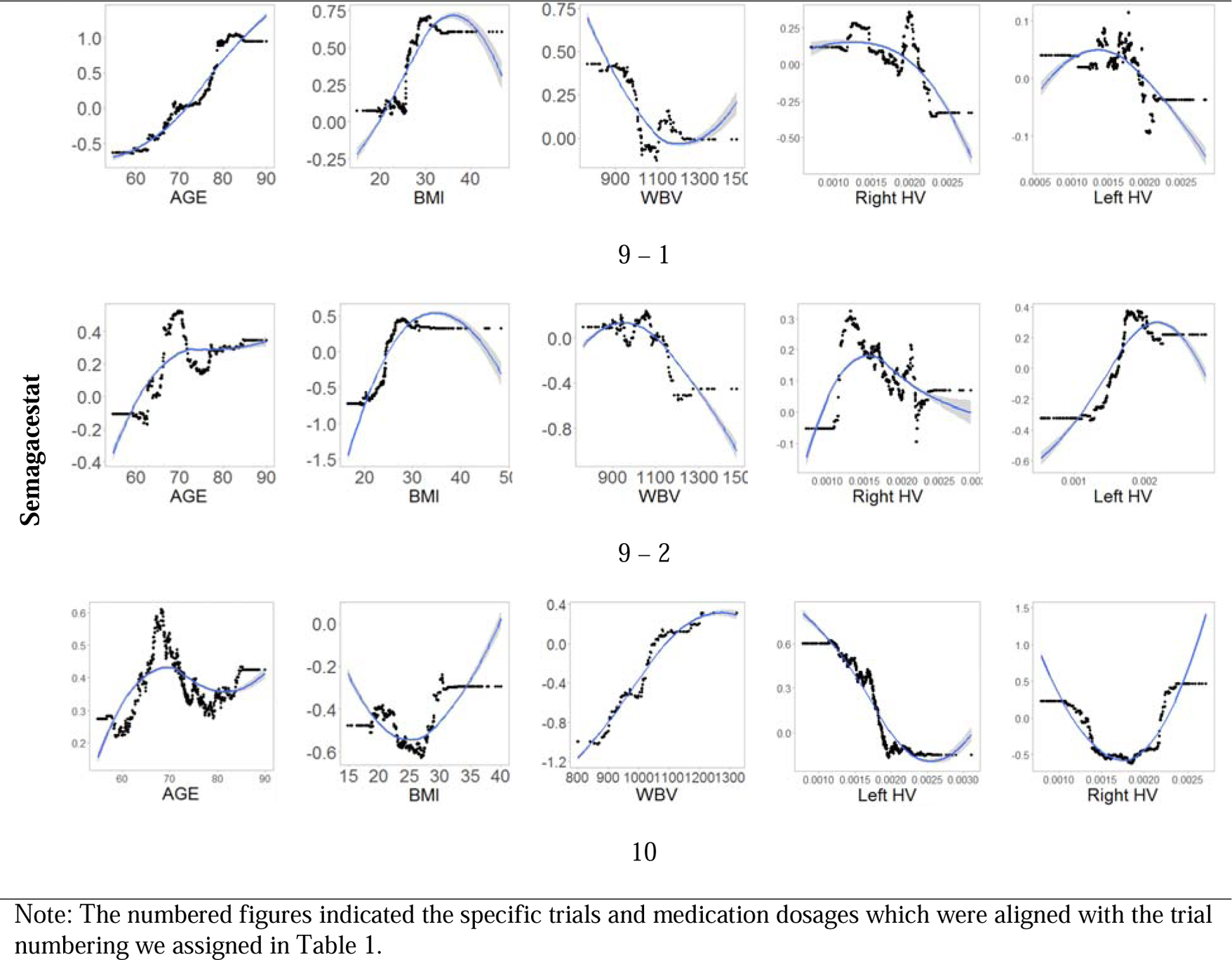
Relationship between CATE and moderators (one moderator vs. CATE)

We focused on features presenting in characteristics of responders within Galantamine trials 5 and 6. Of particular interest in trial 5, there are negative correlations between the CATE and variables such as BMI (<30), ADL total scores (<40), and whole brain volume (<1100 ml). Conversely, a positive correlation was observed between the CATE and variables such as age (>70) and the right hippocampal volume. In trial 6, the associations of CATE with age and ADL total scores (>40) were reversed.

We then shifted to areas manifesting negative estimated CATE in Bapineuzumab and Semagacestat trials. A consistent trend was observed in Bapineuzumab trials: ventricular volume (<0.05) positively associated with the CATE. Additionally, the whole brain volume (<1100ml, in trials 7-1 and 7-2), Aβ_42_ (>6, in trials 7-1, 7-2, and 8), and Aβ_40_ (in trials 7-2 and 8) had positive associations with the CATE, a negative association with the CATE was observed for right hippocampal volume (trial 8), Aβ_42_ (<6, in trials 7-1 and 7-2). In Semagacestat trials 9-1 and 9-2, we observed that age and BMI were positively associated with the CATE. The whole brain volume (<1100 ml) and right hippocampal volume (>0.0015) were negatively associated with the CATE. In trial 10, the relationship between CATE and moderators was fluctuant. The BMI (<25) and right hippocampal volume (<0.0015) were negatively associated with CATE, whereas BMI (>25), right hippocampal volume (>0.002), and whole brain volume were positively associated with CATE.

### Evaluation

We evaluated whether the causal forest accurately estimated CATE and captured the heterogeneity by first comparing subgroups with lower and higher estimated CATE and then using the treatment heterogeneity test. In our setting, patients below the median of the overall range of CATE are likely to benefit from treatment, while those above the median had no response or negative responses to treatments. In addition, we demonstrated that the estimated CATE in the lower and higher regions are separated, indicating that the causal forests captured the heterogeneity of treatment effects (see eTable 1 in the Supplement). We also observed that the trials had varying values of α and β, with some trials showing high significance levels or having values close to 1. For the trials where β had negative values, we also checked the estimated CATE at lower quantile (Q1) and higher quantile (Q3) (See Table 2 and Table 3). The tables suggested that the causal forest estimates performed reasonably well in estimating the CATE in the trials analyzed.

**Table 3.** Coefficients of heterogeneity test.

| of trial | Mean forest prediction $\alpha$ | Differential forest prediction $\beta$ | CATE in Lower ITE region (<median) | CATE in Higher ITE region (>median) | 95% CI |
| --- | --- | --- | --- | --- | --- |
| 1 | 1.03 *** | -1.11 | -11.71 | 1.87 | 13.58 (8.84, 18.34) |
| 2 | 1.00** | -0.21 | -1.40 | 0.29 | 1.69 (1.00, 2.38) |
| 3-1 | 1.02 *** | -1.29 | -5.44 | 0.41 | 5.85 (3.67, 8.03) |
| **3-2** | 1.01 *** | -0.18 | -6.63 | 0.48 | 7.11 (4.94, 9.29) |
| **4-1** | 0.88 * | -0.09 | -4.47 | 1.80 | 6.27 (3.46, 9.08) |
| **4-2** | 1.01 *** | 0.27 | -6.96 | 0.45 | 7.41 (5.37, 9.46) |
| **4-3** | 1.03 *** | 0.47 | -6.34 | 0.22 | 6.56 (4.39, 8.73) |
| **5** | 0.84 | -0.42 | -3.47 | 3.68 | 7.15 (5.93, 8.37) |
| **6** | 1.06 | 0.19 | 3.29 | -3.05 | 6.34 (4.62, 8.06) |
| **7-1** | 1.05 | -0.73 | -7.19 | 6.98 | 14.17 (11.96, 16.37) |
| **7-2** | 0.60 | 0.85 | -6.34 | 6.34 | 12.68 (9.77, 15.58) |
| **8** | 0.98 | 0.52 | 6.93 | 6.76 | 13.69 (11.41, 15.96) |
| **9-1** | 1.25 | -1.32 | -5.63 | 5.47 | 11.10 (9.16, 13.04) |
| **9-2** | 1.20 | -2.88 | -6.74 | 9.8 | 12.98 (10.40, 15.56) |
| **10** | 1.15 | -0.16 | -4.92 | 4.72 | 9.63 (7.66, 11.62) |

Significance codes with p-value: 0 '\*\*\*', 0.001 '\*\*', 0.01 '\*', 0.05 '.', 0.1 '. '. The values of $\alpha$ and $\beta$ for each trial are listed in the table, with different significance codes indicating the level of significance. Negative CATE indicates benefit.

## DISCUSSION

This study aimed to identify important pre-treatment features that moderate treatment effects and thus define subgroups with different treatment effects in Alzheimer’s disease clinical trials. We used causal forest modeling on individual-level patient data from ten randomized clinical trials to estimate CATE, characterize responders and non-responders, and identify potential moderators. The characteristics of non-responders and responders varied considerably across the trials and disease severity stages. This variability suggests that a complex interplay of multiple factors, including BMI, age, amyloid burden, and brain atrophy, might influence responsiveness to Galantamine, Bapineuzumab, and Semagacestat. These findings are valuable as they can help tailor treatment strategies for specific subpopulations within existing developed drugs, thereby reducing costs and reducing exposure to those unlikely to respond. Our method may be a promising tool to inform future clinical trial designs.

Specifically, overweight patients across all dementia stages showed no response to the Galantamine treatment. These heavier persons may have a different metabolism that is related to different responses to treatment. This finding aligns with prior research suggesting a potential link between a higher BMI and reduced reaction to certain AD treatments due to inflammation and vascular issues.^34^ In contrast, we noted that underweight patients with mild to moderate AD at baseline also showed no response to the treatment. The exact mechanism connecting BMI and treatment efficacy remains unclear; it could involve synergistic effects with the patient’s dementia status, cognitive scales, and neuropsychological features at baseline. Intriguingly, our results revealed that older, mild to moderate AD patients and younger MCI patients did not respond to Galantamine. This may be attributed to how Galantamine works. Galantamine enhances the levels of a specific natural substance in the brain required for memory and thought, potentially improving cognitive abilities or decelerating their decline in individuals with AD.^35^ Furthermore, it assists in maintaining ADL performance in AD patients.^36^ Therefore, patients with superior cognitive abilities, greater independence, and possibly younger age may not reap significant benefits from Galantamine treatment.

Clinical trials for Bapineuzumab and Semagacestat were halted due to low efficacy.^9,19^ However, our analysis reveals specific subpopulations of mild to moderate AD patients, identified by distinct Aβ levels and hippocampal volume, who might have responded positively to the treatment. In the Bapineuzumab trials, responders consistently exhibited lower Aβ levels. As a humanized anti-Aβ monoclonal antibody, Bapineuzumab has shown its ability to modify Aβ accumulation.^9^ This may account for lower baseline Aβ levels being more likely to achieve complete amyloid clearance, which helps maintain or improve primary outcomes. Our findings suggest that a subpopulation bearing a lesser amyloid plaque burden is potentially more receptive to Bapineuzumab. In the Semagacestat trials, responders had larger right hippocampal volume and more brain atrophy (smaller whole brain volume) at baseline. This asymmetry in hippocampal volume influencing treatment response is intriguing and suggests that further research into this is important. The underlying mechanism might be tied to the lateralization of brain function and the disease’s differential impact on brain hemispheres.^37^

ApoE genotype didn’t show up in the top important features in our analysis due to the tree-based model favoring continuous variables in node splitting. However, so far, we have been learning that ApoE is an important biological factor affecting AD progression as well as AD treatment efficacy.^38^ Lecanemab, a recently approved therapy for adult AD patients, demonstrates efficacy predominantly among ApoE4 carriers benefit from Lecanemab. ^39^ Similarly, only ApoE4 carries responded to Solanezumab treatment.^40^ Thus, there is a precedent for subsets of AD patients responding differently to treatments. Given this, it is worth employing our methodology to identify responsive subgroups within these promising clinical trials.

### Limitation

Several limitations to our study should be considered when interpreting the results. In contrast to traditional hypothesis testing approaches that pre-specify potential moderators based on biological knowledge, our data-driven strategy may produce moderator combinations without a biological basis, leading to spurious results. Due to the variance term in the splitting criterion, tree-based approaches like the causal forest tend to favor continuous variables for node splitting. Thus, important discrete variables may need to be addressed. Finally, we did not consider differences in follow-up period length when investigating treatment effects; we assume the treatment effect is independent of the follow-up period.

## Data Availability

All data produced in the present study are available upon reasonable request to the authors

## Acknowledgements

The authors would like to thank Christine M. Farrell, Ph.D. for her contribution in securing database access for this study. This publication is based on data contributed by Eli Lilly that has been made available through Vivli, Inc. Vivli has not contributed to or approved, and Vivli and Eli Lilly are not in any way responsible for, the contents of this publication. In addition, this study also used data obtained from the Yale University Open Data Access Project under YODA Project 2020-4323, which has an agreement with JANSSEN RESEARCH & DEVELOPMENT, L.L.C. The interpretation and reporting of research using this data are solely the responsibility of the authors and do not necessarily represent the official views of the Yale University Open Data Access Project or JANSSEN RESEARCH & DEVELOPMENT, L.L.C.

